# Prevalence and associated factors of hypertension in women of reproductive age in Bangladesh based on JNC-8 & AHA-2017 guidelines: Evidence from Bangladesh Demographic & Health Survey 2022

**DOI:** 10.1101/2025.09.16.25335939

**Authors:** Tanvir Ahmed, Imran Hossain Monju, Abdul Hannan Akib, Riya Khulal, Chironjit Das, Mohammad Rashidul Hashan

## Abstract

**Background:** In Bangladesh, using the 2017 ACC/AHA guideline has notably altered both the prevalence estimates and the associated factors compared to JNC 7 guideline. According to these guidelines, we investigated the most-recent data to compare hypertension prevalence and risk factors in women of reproductive age (WRA).

**Methods:** We analyzed data from the nationally representative Bangladesh Demographic and Health Survey 2022. Among eligible WRA, blood pressure was measured three times ≥5 minutes apart with an automated device by trained personnel. The mean of the 2nd and 3rd readings was used. Hypertension was defined based on JNC-8 (SBP ≥140 mmHg or DBP ≥90 mmHg or current antihypertensive use) and AHA-2017 (SBP ≥130 mmHg or DBP ≥80 mmHg) criteria. Prevalence ratios (PRs) were estimated to assess the relative burden of hypertension. Poisson regression models with robust variance were used to identify associated factors.

**Results:** Among 4,814 participants, hypertension was present among 13.6% (655) and 40.8% (1962) according to the JNC-8 criteria and 2017 ACC/AHA guidelines, respectively. The prevalence among rural residents more than doubled (from 16.9%, 95% CI: 15.6–18.4% to 40.2%, 95% CI: 38.3–42.1%). Among women aged 18–29, it increased from 6.4% (95% CI:5.1–7.9%) to 25.8% (95% CI:23.5–28.2%). Similar two- to threefold increases were observed across BMI, education, wealth index, contraceptive use, and comorbidities. In adjusted models, age, obesity, and oral contraceptive use were consistently associated with higher hypertension risk under both JNC-8 and ACC/AHA. Diabetes was associated with higher prevalence only under the ACC/AHA definition. Women in Rajshahi and Sylhet region had higher risk under JNC-8. Across guidelines, obesity showed the strongest association.

**Conclusion:** The prevalence of hypertension among reproductive-aged women in Bangladesh is high, especially under the 2017 ACC/AHA guidelines. Key risk factors include age, obesity, diabetes, and oral contraceptive use, with notable disparities by residence and socioeconomic status.

## Introduction

Hypertension, a significant global health challenge, affects an estimated 1.28 billion adults worldwide [1], with nearly half of those individuals unaware of their condition [2]. The prevalence of hypertension has been rising over the decades, mostly attributed to lifestyle changes, increasing obesity rates, and other socioeconomic factors [3,4]. The Global Burden of Disease (GBD) 2019 report attributes approximately 10.8 million deaths globally to hypertension, contributing to nearly 19% of all cardiovascular-related fatalities [5]. This escalating burden is particularly concerning for reproductive-aged women, as it increases the risk of maternal morbidity, long-term cardiovascular diseases (CVDs), and reduced quality of life [5]. Moreover, untreated hypertension in this population contributes to disability [6] and imposes substantial financial strain on healthcare systems [7], with global healthcare costs associated with hypertension estimated at over $370 billion annually[8].

The burden of hypertension is a major public health concern in developing countries like Bangladesh. Approximately 28.9% of adult women in Bangladesh have elevated blood pressure [9]. According to the 2017–2018 Bangladesh National STEPS Survey, the prevalence among women of reproductive age (WRA) was reported at 17.9% [10]. Similar rising trends were also observed across neighboring countries, e.g., in India, the National Family Health Survey (NFHS-4) revealed a growing prevalence of hypertension among WRA reaching up to 11.6% in some regions [11]. This trend is alarming, as hypertension during pregnancy can result in severe complications, including maternal mortality, one of the leading causes of maternal death in both developed and developing countries [12,13].

Several risk factors have been identified for hypertension among WRA. These include maternal age, pre-pregnancy obesity, excessive weight gain during pregnancy, and gestational diabetes [14–16]. Additionally, socioeconomic factors such as income, education level, and access to health care play a crucial role in shaping the prevalence of hypertension in this population [17]. Lifestyle-related factors, such as physical activity, dietary habits, and the use of tobacco or alcohol, are also identified as significant contributors [18–21]. In low-resource settings, women may have limited access to routine screening, contributing to the underreporting of hypertension cases [22].

“The Eighth Joint National Committee on Prevention, Detection, Evaluation and Treatment of High Blood Pressure 2003 guidelines (JNC-8)”, and “2017 American College of Cardiology/American Heart Association (2017 ACC/AHA) Guideline for the Prevention, Detection, Evaluation, and Management of High Blood Pressure in Adults” provide evidence based method to classify hypertension [23,24]. The JNC-8 guidelines classify hypertension at a blood pressure of ≥140/90 mmHg for most adults. In contrast, the 2017 AHA guidelines lowered the threshold to ≥130/80 mmHg, redefining hypertension and encouraging earlier intervention. According to clinical trials, this lower cut-off significantly improves patient outcomes [25–28]. Following the new ACC/AHA guidelines, previously undiagnosed patients or prehypertensive individuals are diagnosed as hypertensive [29–41]. Using cut-off from both JNC-8 and ACC/AHA 2017 guidelines, a cross-sectional analysis with the most recent data can provide a snapshot of the current situation of hypertension among non-pregnant reproductive-aged women in Bangladesh. Associations between various factors, such as socioeconomic status and lifestyle behaviors, can be assessed.

Thus, we aim to identify this using the latest Bangladesh Demographic and Health Survey (BDHS) 2022 data to determine the prevalence and associated factors of hypertension among non-pregnant reproductive-aged women in Bangladesh using JNC-8 and ACC/AHA 2017 guideline criteria.

## Materials & Methods

### Data Sources

The study analyzed data from the 2022 Bangladesh Demographic and Health Survey (BDHS) latest dataset[44]. The survey was conducted from January to December 2022 under the supervision of the National Institute of Population Research and Training (NIPORT), with support from the Ministry of Health and Family Welfare (MoHFW) and the United States Agency for International Development (USAID). The DHS data sets are globally recognized as nationally representative and comparable surveys. The survey is conducted in over 90 countries worldwide. To organize the survey effectively, the program possesses standardized manuals, data collection methods, and variables. The BDHS’s primary goal was to collect data on a range of demographic, health, and socio-economic indicators. Indicators of non-communicable diseases (NCDs) such as hypertension and diabetes are assessed. Special emphasis was given to indicators of maternal and child health, such as antenatal care, unmet need for family planning, intimate partner violence, under-5 mortality, etc.

### Study Population and Survey Design

The sampling frame for the 2022 BDHS was derived from the 2011 Population and Housing Census conducted by the Bangladesh Bureau of Statistics[44] . The survey utilized a two-stage stratified cluster sampling design. In the first stage, 675 enumeration areas (EAs) were selected with probability proportional to size. The survey covered both rural and urban areas, ensuring proportional representation from all eight administrative divisions. In the second stage, systematic random sampling was employed to select households from each EA. The survey team collected comprehensive demographic, health, and biomarker data, including blood pressure measurements, to assess hypertension prevalence among women. Biomarker data were collected in a subsample of households in the 2022 BDHS. The biomarkers collected included anthropometry (height and weight), blood pressure, and blood glucose measurements. In one-fourth of the households randomly selected for the 2022 BDHS survey sample, all women and men aged 18 and older were eligible to participate in biomarker measurements, including blood pressure measurement, fasting plasma blood glucose testing, and height and weight measurements. 8,156 women and 6,853 men aged 18 and older were eligible for blood pressure and blood glucose measurements. Among those eligible, 95% of women and 91% of men were measured for blood pressure [44]. We restricted our analysis to women of reproductive age because, in the BDHS 2022, height, weight, and biomarker data such as blood pressure and hemoglobin were not collected for men who were interviewed. Due to this subsampling in the survey design, men’s anthropometric and biomarker data cannot be linked with variables from the men’s dataset. For this study, we merged the Individual Recode (IR) dataset with the Household Member Recode (PR) dataset.

Out of 5,098 reproductive-aged women whose blood pressure was measured, we excluded 284 pregnant women. Data from the remaining 4814 non-pregnant women of reproductive age were analyzed for the prevalence of hypertension (Fig 1). When exploring associated factors, due to missing values in some of the variables, the total weighted number of observations was 4680 for our analysis.

**Figure 1:** Flow chart of sampling method and sample size.

### Outcome Variable

The primary outcome variable in this study was hypertension. Blood pressure was measured using the Multi-User Upper Arm Blood Pressure Monitor UA-767F/FAC. Data collectors had three monitors with required cuff sizes (medium, small, and extra-large). Three blood pressure readings were taken at intervals of at least five minutes. Based on the WHO guidelines, the average of the second and third readings was used to classify hypertension. As per the guideline, systolic blood pressure (SBP) ≥ 140 mmHg or diastolic blood pressure (DBP) ≥ 90 mmHg constitutes hypertension[45]. We classified hypertension based on JNC-8 (SBP ≥140 mmHg or DBP ≥90 mmHg or current antihypertensive use) and AHA-2017 (SBP ≥130 mmHg or DBP ≥80 mmHg) criteria[33,45].

### Explanatory Variables

Explanatory variables were selected from existing literature on hypertension risk factors in low- and middle-income countries (LMICs)[9,41,46]. Age of the respondent was categorized into four groups (15-24, 25-34, 35-44, 45-49 years). Education is grouped into four categories (no education, primary, secondary, and higher). Other variables included were wealth index (poorest, poorer, middle, richer, richest); occupation (working, not working); frequency of birth in the last five years (no, single, multiple), administrative division (Barisal, Chittagong, Dhaka, Khulna, Mymensingh, Rajshahi, Rangpur, and Sylhet), place of residence (urban, rural); contraceptive use (not using, oral pills, other methods); diabetes (absent, present); depression (absent, present); anxiety (absent, present). We defined diabetes in WRA with fasting plasma glucose values of 7.0 mmol/L and above, depression as WRA having a PHQ-9 score of 10 or higher [47] and anxiety as WRA with a GAD-7 score is 6 or higher[48]. Although depression and anxiety were not clinically diagnosed, the use of validated screening scales is widely accepted in population-based surveys to estimate symptom burden and identify high-risk groups. Body Mass Index (BMI) is classified using the WHO recommendations for Asians. BMI of WRA is classified into underweight (<18.5 kg/m²), normal (18.5-22.9 kg/m²), overweight (23.0-24.9 kg/m²), and obese (≥25.0 kg/m²)[23,49,50].

### Statistical Analysis

Descriptive statistics of the selected variables were performed and presented by frequency distribution. We assessed the normality of continuous variables based on their distribution and non-normally distributed data were presented using medians and interquartile ranges (IQRs). The prevalence of hypertension was estimated using weighted proportions to account for the complex survey design of the BDHS. We used the svy: proportion command in STATA version 17.0. Estimates were reported with 95% confidence intervals. In the analysis of data from cross-sectional studies, the Poisson models with robust variance are better alternatives than logistic regression [51]. The log-binomial regression model produces unbiased PR estimates but may present convergence difficulties when the outcome is very prevalent, and the confounding variable is continuous[52]. Considering all this, we performed Poisson models with robust variance to identify significant factors associated with hypertension. Prevalence ratios (PRs) were estimated to identify factors associated with hypertension. The final multivariate model included only those variables that demonstrated statistical significance (p < 0.05) in univariate analyses. All statistical analysis was performed using STATA version 17.0 [53].

### Ethical Considerations

The study was based on secondary data from the BDHS, which is publicly accessible. The ethical guidelines and protocols adhered to by the BDHS are documented in the 2022 BDHS report [44]. Ethical approval for the BDHS was obtained from the Institutional Review Board (IRB) of ICF International and the Ethical Review Committee of NIPORT, Bangladesh. All respondents provided informed consent prior to participation, and all data were anonymized prior to public release. No additional data collection or direct interaction with human or animal subjects was conducted for this analysis. Therefore, additional ethical approval was not required. Approval of using the dataset was obtained from the DHS program. The study adhered to the principles of the Declaration of Helsinki.

## Results

### Background Characteristics of the Participants

Among 4,814 participants, hypertension was present among 13.6% (n=655) and 40.8% (n=1962) according to the JNC-8 criteria and 2017 ACC/AHA guidelines, respectively. The median (IQR) age of the participants was 32.0 years (25.0–40.0), while the highest proportion belonged to 18-29 years of age (36.9%), and more than two-thirds (71.6%) of them resided in rural areas. Most participants (88.3%) had measured their blood pressure at least once in their lifetime, while only 10.3% (494) were aware of their hypertension status. The median BMI was 23.5 (20.7–26.5), and 15.1% (706) had no formal education. Among the other medical conditions, diabetes (14.2%), depression (6.7%), and anxiety (21.2%) were also present among study respondents (Table 1).

**Table 1:** Distribution of respondents by background characteristics.

| Background characteristics | BDHS 2022 |  |  |
| --- | --- | --- | --- |
|  | All participants<br>(N = 4,814),<br>n (%) | Hypertensive<br>participants per JNC-8,<br>N= 655,<br>n (%) | Hypertensive<br>participants per<br>2017 ACC/AHA,<br>N = 1962<br>n (%) |
| SBP, median (IQR), (mmHg) | 111 (103–121) | 141 (150–128) | 133 (122–147) |
| DBP, median (IQR), (mmHg) | 77 (71–83) | 91 (94–99) | 85 (82–90) |
| Ever measured BP | 4250 (88.3) | 620 (94.5) | 1796 (91.5) |
| Know about the hypertension status | 494 (10.27) | 263 (40.1) | 469 (23.9) |
| **Administrative divisions** |  |  |  |
| Barisal | 305 (6.52) | 86 (9.6) | 247 (12.33) |
| Chittagong | 891 (19.04) | 141 (15.74) | 289 (14.42) |
| Dhaka | 1113 (23.79) | 114 (12.72) | 245 (12.23) |
| Khulna | 564 (12.05) | 118 (13.17) | 277 (13.82) |
| Mymensingh | 369 (7.89) | 86 (9.6) | 208 (10.38) |
| Rajshahi | 622 (13.29) | 123 (13.73) | 285 (14.22) |
| Rangpur | 530 (11.33) | 99 (11.05) | 216 (10.78) |
| Sylhet | 285 (6.1) | 129 (14.40) | 237 (11.83) |
| **Place of residence** |  |  |  |
| Urban | 1327 (28.36) | 367 (40.96) | 741 (37) |
| Rural | 3353 (71.64) | 529 (59.04) | 1263 (63) |
| **Age of the participants (years)** |  |  |  |
| Median (IQR) | 32.0 (25.0–40.0) | 39.0 (33.0–44.0) | 36.0 (30.0–43.0) |
| 18–29 | 1727 (36.9) | 121 (13.50) | 470 (23.45) |
| 30–39 | 1694 (36.2) | 342 (38.17) | 789 (39.37) |
| 40–49 | 1259 (26.9) | 433 (48.33) | 745 (37.18) |
| **BMI (body mass index) (kg/m<sup>2</sup>)** |  |  |  |
| Median (IQR) | 23.51 (20.69–26.48) | 25.59 (23.14–28.73) | 25.1 (22.27–28) |
| Underweight | 483 (10.33) | 33 (3.68) | 101 (5.04) |
| Normal | 1582 (33.79) | 179 (19.98) | 501 (25.0) |
| Overweight | 885 (18.9) | 180 (20.09) | 382 (19.06) |
| Obesity | 1730 (4.1) | 504 (56.25) | 1020 (50.9) |
| **Educational Status** |  |  |  |
| No education | 706 (15.08) | 145 (22.69) | 346 (18.12) |
| Primary | 1271 (27.15) | 190 (29.66) | 562 (29.43) |
| Secondary | 2089 (44.64) | 245 (38.42) | 790 (41.39) |
| Higher | 614 (13.13) | 104 (11.6) | 211 (11.06) |
| **Occupational Status** |  |  |  |
| Not working | 2788 (59.58) | 497 (57.63) | 1110 (57.43) |
| Working | 1892 (40.42) | 365 (42.37) | 823 (42.57) |
| **Wealth Status** |  |  |  |
| Poorest | 830 (19.4) | 125 (14.55) | 336 (17.4) |
| Poorer | 958 (19.7) | 155 (18.02) | 369 (19.11) |
| Middle | 937 (20.6) | 162 (18.82) | 384 (19.88) |
| Richer | 996 (21.27) | 216 (25.09) | 414 (21.41) |
| Richest | 959 (20.5) | 203 (23.52) | 429 (22.20) |
| **Contraceptive use history** |  |  |  |
| No | 1588 (33.92) | 328 (38.08) | 685 (35.42) |
| Oral Pill | 1295 (27.67) | 257 (29.81) | 557 (28.80) |
| Other methods | 1797 (38.41) | 277 (32.11) | 692 (35.79) |
| **Birth in the last five years** |  |  |  |
| No birth | 2845 (60.79) | 688 (79.74) | 1385 (71.6) |
| Single | 1584 (33.85) | 157 (18.26) | 488 (25.25) |
| Multiple | 251 (5.36) | 17 (2) | 61 (3.15) |
| **Diabetes** |  |  |  |
| Absent | 4014 (85.78) | 651 (75.55) | 1585 (81.97) |
| Present | 666 (14.22) | 211 (24.45) | 349 (18.03) |
| **Depression** |  |  |  |
| Absent | 4367 (93.32) | 799 (92.7) | 1798 (93) |
| Present | 313 (6.68) | 63 (7.3) | 136 (7) |
| **Anxiety** |  |  |  |
| Absent | 3686 (78.76) | 651 (75.55) | 1479 (76.5) |
| Present | 994 (21.24) | 211 (24.45) | 454 (23.5) |

### Prevalence and Distribution of Hypertension Burden Using JNC-8 and AHA-2017 Guidelines

According to the JNC-8 criteria, the prevalence of hypertension in the Barishal division was 15.9% (95% CI: 12.4–20%), while according to ACC/AHA criteria, it was 46.9% (95% CI: 41.7–52.2%). This shows almost a threefold increase in prevalence with later guidelines. With other divisions, the prevalence has increased at least twice in the ACC/AHA criteria. Prevalence among rural residents was more than double according to ACC/AHA criteria (40.2%, 95% CI: 38.3–42.1%) than the JNC-8(16.9%, 95% CI: 15.6–18.4%). After using the ACC/AHA method for defining hypertension, the prevalence among 18-29-years women increased from 6.4% (95% CI: 5.1–7.9%) to 25.8% (95% CI: 23.5–28.2%). A similar two to three-fold increase in prevalence was also seen for other variables such as BMI, education level, wealth index, contraceptive use, and other comorbid conditions. (Table 2)

**Table 2:** Weighted prevalence of hypertension according to selected characteristics.

| Explanatory variables | Prevalence of hypertension per JNC-8, prevalence (95% CI) | Prevalence of hypertension per 2017 ACC/AHA, prevalence (95% CI) | Difference in prevalence |
| --- | --- | --- | --- |
| **Administrative Divisions** |  |  |  |
| Barisal | 15.9 (12.4–20) | 46.9 (41.7–52.2) | 31.0 |
| Chittagong | 18.8 (16.0–22.0) | 38.4 (34.3–42.8) | 19.6 |
| Dhaka | 18.1 (15.1–21.4) | 38 (33.9–42.2) | 19.9 |
| Khulna | 18.9 (16.1–22.2) | 45.7 (42–49.4) | 26.8 |
| Mymensingh | 16.3 (13.2–19.8) | 40.3 (36.1–44.7) | 24 |
| Rajshahi | 20.3 (16.8–24.2) | 47.8 (43–52.9) | 27.5 |
| Rangpur | 15.7 (12.8–19) | 36.1 (32.1–40.3) | 20.4 |
| Sylhet | 24.3 (20.5–28.6) | 45.5 (40.8–50.3) | 21.2 |
| **Place of Residence** |  |  |  |
| Urban | 22.2 (19.7–25) | 44.0 (40.8–47.7) | 21.8 |
| Rural | 16.9 (15.6–18.4) | 40.2 (38.3–42.1) | 23.3 |
| **Age Groups** |  |  |  |
| 18–29 | 6.4 (5.1–7.9) | 25.8 (23.5–28.2) | 19.4 |
| 30–39 | 19.7 (17.6–22) | 45.6 (42.9–48.4) | 25.9 |
| 40–49 | 33.3 (30.5–36.1) | 56.9 (53.7–60) | 23.6 |
| **BMI** |  |  |  |
| Underweight | 6.8 (4.8–9.7) | 19.5 (16–23.5) | 12.7 |
| Normal | 10.6 (9.1–12.3) | 30.4 (28–32.9) | 19.8 |
| Overweight | 19.8 (17–22.8) | 42.3 (38.7–45.9) | 22.5 |
| Obesity | 28.1 (25.8–30.6) | 56.9 (54.1–59.7) | 28.8 |
| **Education Level** |  |  |  |
| No education | 24.7 (21.2–28.6) | 49.6 (45.4–53.9) | 24.9 |
| Primary | 20.5 (18.1–23.2) | 44.6 (41.6–47.7) | 24.1 |
| Secondary | 16.3 (14.6–18.1) | 38.4 (36–40.8) | 22.1 |
| Higher | 14.3 (11.5–17.7) | 34.9 (30.9–39.1) | 20.6 |
| **Occupation** |  |  |  |
| Not working | 17.8 (16.3–19.5) | 39.8 (37.7–42) | 22.0 |
| Working | 19.3 (17.4–21.4) | 43.5 (41–46) | 24.2 |
| **Wealth Index** |  |  |  |
| Poorest | 15.1 (12.6–18) | 40.5 (36.9–44.2) | 25.4 |
| Poorer | 16.2 (13.9–18.8) | 38.6 (35.2–42.1) | 22.4 |
| Middle | 17.3 (14.7–20.3) | 41 (37.6–44.5) | 23.7 |
| Richer | 21.7 (19.1–24.7) | 41.6 (38.2–45.1) | 19.9 |
| Richest | 21.1 (18.6–23.9) | 44.8 (41.1–48.5) | 23.7 |
| **Contraceptive Use** |  |  |  |
| Not using | 20.7 (18.6–22.9) | 43.1 (40.2–46.1) | 22.4 |
| Oral pill | 19.9 (17.4–22.5) | 43 (40.1–45.9) | 23.1 |
| Other methods | 15.4 (13.7–17.3) | 38.5 (35.9–41.1) | 23.1 |
| **Frequency of birth in the last five years** |  |  |  |
| No birth | 24.2 (22.4–26.1) | 48.7 (46.4–51) | 24.5 |
| Single | 9.9 (8.4-11.8) | 30.8 (28.3-33.5) | 20.9 |
| Multiple | 6.9 (4.3-10.9) | 24.3 (19.5-29.8) | 17.4 |
| **Diabetes** |  |  |  |
| Absent | 16.2 (15-17.5) | 39.5 (37.7-41.3) | 23.3 |
| Present | 31.7 (27.6-36.1) | 52.4 (47.8-56.9) | 20.7 |
| **Depression** |  |  |  |
| Absent | 18.3 (17-19.7) | 41.2 (39.4-42.9) | 22.9 |
| Present | 20.1 (15.8-25.4) | 43.4 (37.8-49.1) | 23.3 |
| **Anxiety** |  |  |  |
| Absent | 17.7 (16.3-19.1) | 40.1 (38.3-42) | 22.4 |
| Present | 21.3 (18.5-24.3) | 45.7 (42.1-49.3) | 24.4 |
| *CI* confidence interval, *BMI* body mass index. |  |  |  |

### Risk Factors for Hypertension

After adjusting for covariates, the hypertensive risk remained significant among respondents from Rajshahi (aPR: 1.48, 95% CI: 1.06–2.06, p = 0.021) and Sylhet (aPR: 1.50, 95% CI: 1.08–2.07, p = 0.015) in the JNC-8 guideline while Chattogram (aPR: 0.78, 95% CI: 0.68–0.90, p = 0.001) and Dhaka (aPR: 0.75, 95% CI: 0.65–0.88, p = <0.001) divisions had lower risk of hypertension compared to Barishal. Hypertension prevalence increased with age across both guidelines. In the JNC-8 criteria, individuals aged 30-39 (aPR: 2.40, 95% CI: 1.78–3.24, p = <0.001) and aged 40- 49 (aPR: 3.68, 95% CI: 2.69–5.03, p = <0.001) had a higher risk of hypertension and under the ACC/AHA guidelines, 30–39 age groups (aPR: 1.50, 95% CI: 1.33–1.68, p = <0.001) and 40-49 age groups (aPR: 1.76, 95% CI: 1.55–2.01, p = <0.001) had also at higher risk of having hypertension. BMI was strongly associated with an increased risk of hypertension. Obesity had the strongest association with hypertension under both guidelines. Under JNC-8, the prevalence ratio for obesity was significant after adjustment (aPR: 3.56, 95% CI: 2.3–5.52, p = <0.001). Similarly, the ACC/AHA2017 criteria showed that obesity was associated with a threefold increase in hypertension prevalence (aPR: 2.70, 95% CI: 2.21–3.30, p = <0.001). Oral pill users had a higher prevalence under JNC-8 (aPR: 1.29, 95% CI: 1.07–1.57, p = 0.009) and AHA 2017 (aPR: 1.10, 95% CI: 1.01–1.21, p = 0.036). However, other contraceptive methods were associated with a lower prevalence under both guidelines. According to the ACC/AHA 2017 guidelines, diabetic individuals had a significant risk of hypertension (aPR: 1.10, 95% CI: 1.01–1.21, p = 0.031) (Table 3)

**Table 3:**
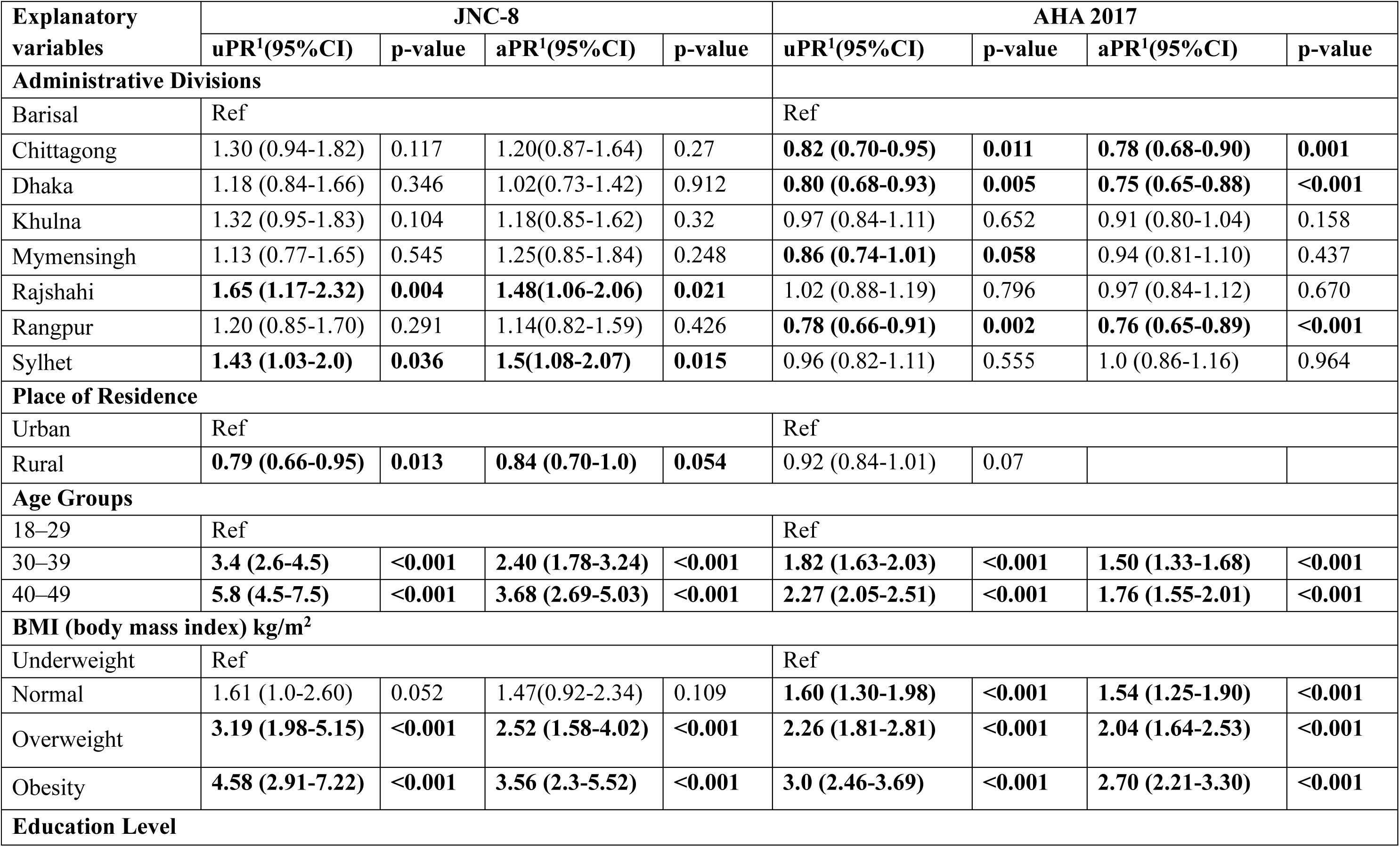
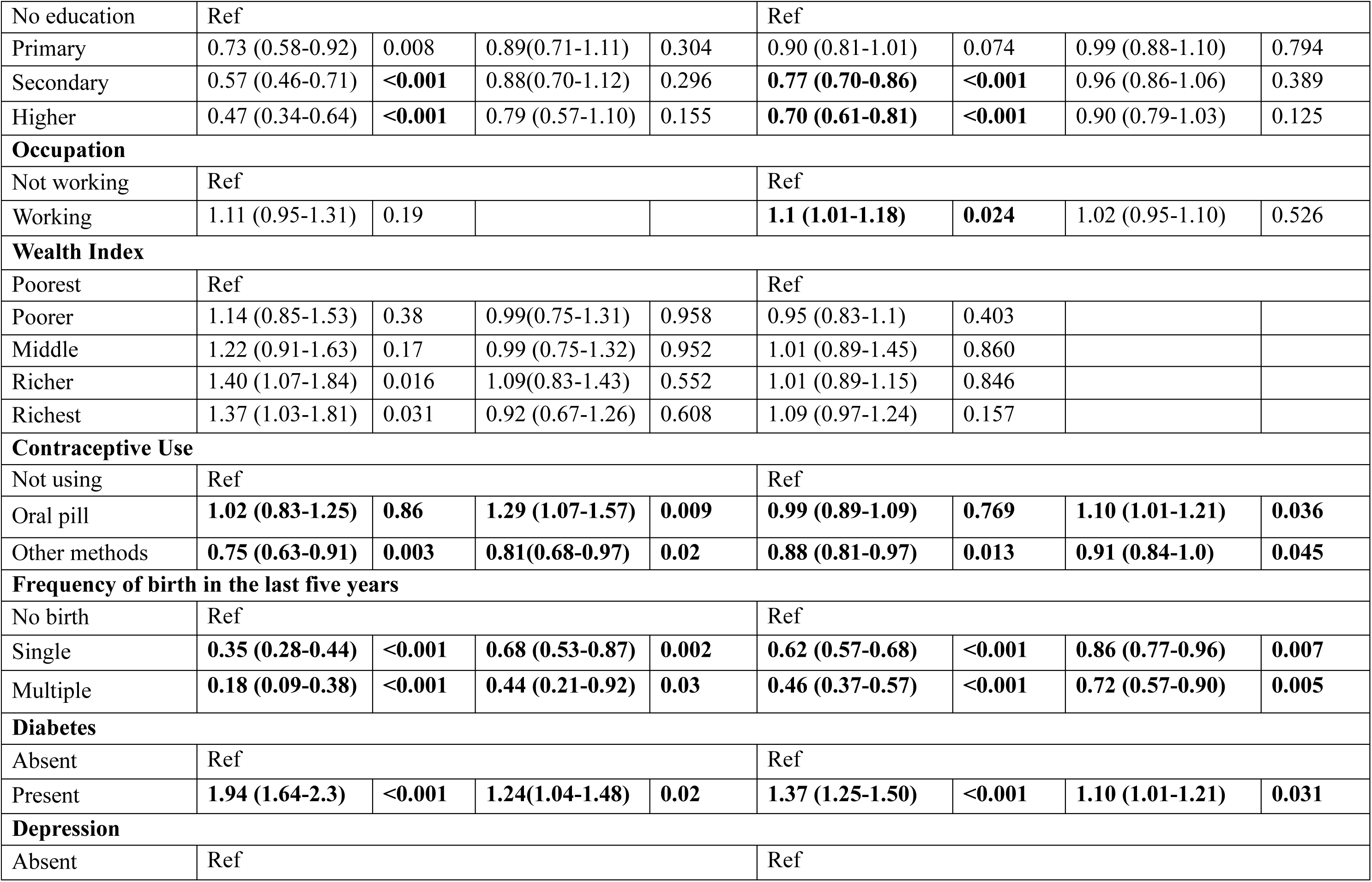
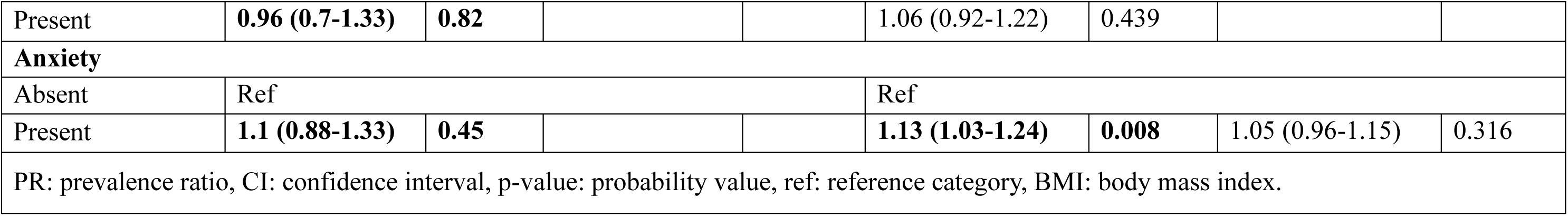
Factors associated with hypertension according to selected characteristics.

## Discussion

Our findings from the BDHS 2022 data demonstrate a significant prevalence of hypertension among non-pregnant women of reproductive age in Bangladesh. 2017 ACC/AHA guidelines estimated prevalence is three times more than JNC-8 guidelines. The prevalence of hypertension was higher among urban people, obese, higher age group, those who use contraceptive, and among people with diabetes. [54].

Following the JNC-8 guidelines, one in every eight non-pregnant women were classified as hypertensive, whereas the 2017 ACC/AHA guidelines provide a rise in prevalence of giving two in every five having hypertension. This discrepancy is not new and reflects the global debate on blood pressure thresholds and their implications for public health [55]. A similar trend has been observed in South Asia, where a shift towards higher hypertension prevalence under the ACC/AHA guidelines has been documented. Previously, the prevalence of hypertension among Bangladeshi adults increased to 50.5% when applying the ACC/AHA guidelines, compared to 24% under the JNC-8 criteria [46]. With recent BDHS data, the discrepancy in prevalence even increased, which warrants more attention on early intervention. The more stringent ACC/AHA criteria may prompt earlier detection and intervention, thereby reducing the risk of long-term cardiovascular complications [56].

The finding of a higher prevalence of hypertension in urban regions compared to rural areas, under both the JNC-8 (40.96% vs. 59.04%) and ACC/AHA (37% vs. 63%) guidelines, is consistent with existing literature [38]. We estimated prevalence ratios (PRs) using Poisson regression with robust variance to identify factors associated with hypertension. Older age, higher BMI, urban residence, and diabetes were associated with hypertension. The distribution of risk groups remained consistent across both guidelines. Urbanization in Bangladesh has been linked to lifestyle changes, including increased consumption of processed foods, physical inactivity, and higher levels of stress, all of which contribute to elevated blood pressure [57]. Studies from other low- and middle-income countries (LMICs) have similarly reported a higher hypertension prevalence in urban settings, highlighting the need for targeted interventions in urban populations[58]. The significant urban-rural disparity emphasizes the need for differentiated public health strategies that address both urban and rural lifestyle risk factors. Rural women, particularly those from marginalized communities, face unique challenges such as limited healthcare access [59] lower health literacy [60,61] and cultural barriers to healthcare-seeking behavior [62]. Public health interventions in rural Bangladesh can be included in community health worker program to improve health literacy and access to preventive care, as well as culturally tailored approaches in addressing lifestyle risk factors [63].

One of the most striking findings of this study is the strong association between obesity and hypertension. Obesity, defined as a BMI ≥25.0 kg/m², was highly correlated with hypertension under both JNC-8 (56.25%) and ACC/AHA (50.9%) guidelines. This is consistent with evidence from other studies in South Asia that the odds of HTN were higher among obese women[63,64]. The underlying mechanisms, such as increased vascular resistance and activation of the sympathetic nervous system, are well-documented [64]. However, the relationship between obesity and hypertension is likely multifactorial, with metabolic dysregulation, inflammation, and hormonal imbalances also playing critical roles [65]. In Bangladesh, the rising prevalence of obesity, particularly in urban settings, necessitates urgent public health interventions to promote healthy diets and physical activity, especially among reproductive-aged women[66]. . Additionally, the impact of obesity on younger vs. older reproductive-aged women remains underexplored. Future research can be delved into these mechanistic differences, particularly concerning hormonal changes during pregnancy, postpartum, and menopause. Age also emerged as a critical factor in hypertension prevalence, with women aged 40–49 years showing significantly higher (33.3% under JNC-8 and 56.9% under ACC/AHA criteria). This pattern aligns with global findings that blood pressure increases with age due to arterial stiffening and other physiological changes[67]. The increased risk among older reproductive-aged women is of particular concern given the potential implications for maternal health, such as the risk of preeclampsia and other pregnancy-related complications (Nayab et al., 2022). Early screening and management of hypertension in this age group are essential for mitigating adverse maternal and cardiovascular outcomes.

Contraceptive use, particularly the use of oral contraceptive pills, was another important factor associated with hypertension in this study. Women using oral contraceptives had a higher prevalence of hypertension (29.81% under JNC-8 and 28.80% under ACC/AHA), which is consistent with previous studies that have identified hormonal contraceptives as a risk factor for elevated blood pressure [68]. The mechanisms behind this association may include estrogen’s effects on vascular function and fluid retention [69]. Given the widespread use of oral contraceptives among women of reproductive age, regular blood pressure monitoring is essential, and healthcare providers can consider alternative contraceptive methods for women at high risk of hypertension. The strong association between diabetes and hypertension, as observed in this study, mirrors global evidence on the comorbidity of these two conditions. Diabetic women in this sample had nearly double the prevalence of hypertension compared to non-diabetic women. This finding is in line with the pathophysiological overlap between diabetes and hypertension, such as insulin resistance and endothelial dysfunction, which jointly contribute to cardiovascular risk [70]. The coexistence of diabetes and hypertension greatly increases the risk of cardiovascular diseases (CVDs) and demands a comprehensive approach to non-communicable disease (NCD) management that includes regular monitoring, patient education, and lifestyle modification programs. With the rising prevalence of diabetes in Bangladesh, particularly in urban areas, integrated public health interventions targeting both diabetes and hypertension are essential for reducing the overall burden of CVD[65].

The urban-rural disparity in hypertension prevalence highlights the need for tailored interventions. Targeted public health policies should address these disparities by focusing on prevention, early detection, and context-specific treatment strategies. Our study findings show that women with advanced age were at risk of hypertension or elevated blood pressure according to both the 2017 ACC/AHA and JNC-8 classifications. The association of hypertension with obesity, contraceptive use, and diabetes indicates the requirement for active lifestyles and healthy dietary habits and cautious use of contraceptives, and optimal control of diabetes. Our study did not find any association between hypertension and wealth index, education level, depression and anxiety. Public health initiatives need to effectively respond to this growing concern. To better manage the hypertension burden, it is crucial to assess its prevalence using the diagnostic thresholds that early detect hypertension and include most of the patients at risk. This approach may help mitigate the risk of escalating cardiovascular diseases. The insights from such assessments could support future research and assist policymakers in developing targeted programs and policies to control and prevent hypertension, ultimately addressing this significant public health issue.

Our study has some strengths and limitations. A key strength lies in the generalizability of its findings to the broader Bangladeshi population, as the analysis was based on nationally representative data encompassing all administrative divisions. Furthermore, the application of appropriate statistical techniques to estimate the weighted prevalence of hypertension enhances the study’s methodological rigor. In our study, we used JNC-8 for comparison rather than JNC-7, as JNC-8 used a more rigorous evidence-based approach, simplifying treatment goals across populations, allowing broader first-line drug choices, omitting definitions of pre-hypertension, and introducing a formal grading system for its recommendations However, certain limitations should be noted. The cross-sectional design precludes the establishment of causal relationships. Additionally, blood pressure was measured three times within a single day, whereas both international guidelines recommend repeated longitudinal measurements for accurate hypertension diagnosis. This may introduce measurement bias. Diagnostic bias was also possible as the data collectors used an automated device for blood pressure assessment, although standard recommendations advise the use of a sphygmomanometer for such measurements. Again, depression and anxiety were assessed using screening scales rather than clinical diagnosis. Without clinical evaluation, this cannot establish a definitive diagnosis. As a result, both recall bias and diagnostic bias may have influenced our findings.

## Conclusion

The prevalence of hypertension among reproductive-aged women in Bangladesh, particularly under the 2017 ACC/AHA guidelines, is notably high. Key risk factors, including age, BMI, diabetes, and contraceptive use, have been identified, alongside significant urban-rural and socioeconomic disparities. These findings highlight the urgent need for targeted public health interventions focused on early detection, lifestyle modification, and, especially for high-risk groups such as older women, diabetics, and oral contraceptive users.

## Data Availability

The data used for this study were obtained from the publicly available Demographic and Health Surveys (DHS) Program database (https://dhsprogram.com).

## Acknowledgements

We gratefully acknowledge the Demographic and Health Surveys (DHS) Program, the National Institute of Population Research and Training (NIPORT), and the Bangladesh Bureau of Statistics (BBS). We also thank the survey participants and field staff for their valuable contributions.

